# Model-based estimates of deaths averted and cost per life saved by scaling-up mRNA COVID-19 vaccination in low and lower-middle income countries in the COVID-19 Omicron variant era

**DOI:** 10.1101/2022.02.08.22270465

**Authors:** Alexandra Savinkina, Alyssa Bilinski, Meagan C. Fitzpatrick, A. David Paltiel, Zain Rizvi, Joshua A. Salomon, Tommy Thornhill, Gregg Gonsalves

## Abstract

**Background:** While almost 60% of the world has received at least one dose of COVID-19 vaccine, the global distribution of vaccination has not been equitable. Only 4% of the population of low-income countries has received a full primary vaccine series, compared to over 70% of the population of high-income nations.

**Methods:** We used economic and epidemiologic models, parameterized with public data on global vaccination and COVID-19 deaths, to estimate the potential benefits of scaling up vaccination programs in low and lower-middle income countries (LIC/LMIC) in 2022 in the context of global spread of the Omicron variant of SARS-CoV2. Outcomes were expressed as number of avertable deaths through vaccination, costs of scale-up, and cost per death averted. We conducted sensitivity analyses over a wide range of parameter estimates to account for uncertainty around key inputs.

**Findings:** Global scale up of vaccination to provide two doses of mRNA vaccine to everyone in LIC/LMIC would cost $35.5 billion and avert 1.3 million deaths from COVID-19, at a cost of $26,900 per death averted. Scaling up vaccination to provide three doses of mRNA vaccine to everyone in LIC/LMIC would cost $61.2 billion and avert 1.5 million deaths from COVID-19 at a cost of $40,800 per death averted. Lower estimated infection fatality ratios, higher cost-per-dose, and lower vaccine effectiveness or uptake lead to higher cost-per-death averted estimates in the analysis.

**Interpretation:** Scaling up COVID-19 global vaccination would avert millions of COVID-19 deaths and represents a reasonable investment in the context of the value of a statistical life (VSL). Given the magnitude of expected mortality facing LIC/LMIC without vaccination, this effort should be an urgent priority.

## Introduction

As of January 7 2022, 59% of the global population had received at least one dose of a COVID-19 vaccine, 50% had received a full primary vaccine course, and 7% had received an additional booster. In low-income countries,^1^ only 9% of people had received any dose of vaccine, and only 4% had received two doses.^2^ The barriers to global access to COVID-19 vaccines have been well-described. Many have emphasized the need to scale-up mRNA vaccine production to remedy these, including the transfer of technology for vaccine production to regional hubs to scale-up supply more quickly as recommend by the World Health Organization.^3,4^ Beyond humanitarian implications, lingering inequities in access to COVID-19 vaccines will delay the worldwide recovery from the pandemic and continue to fuel the rise of new variants as the virus spreads in many parts of the globe.^5^

Because the most dire complications of COVID-19 occur in older people and the age structure in many developing countries skews relatively young, it was initially assumed that the mortality burden in low (LIC) and lower-middle income countries (LMIC) from COVID-19 would be less severe.^6^ However, deaths in LIC/LMIC have been growing as the pandemic has continued, likely due in part to underlying comorbidities.^7^ LIC/LMIC now account for over 50% of the global mortality burden with some researchers suggesting they bear 80% of global COVID-19 mortality when accounting for underreporting of deaths in these regions.^8^ Indeed, due to under-reporting, the recorded number of infections and deaths are likely gross under-estimates of the true burden of the pandemic in LIC/LMICs.^9,10^ Model-based excess mortality estimates suggest that COVID-19-related mortality might be up to 35 times higher than reported figures in these settings. ^8,11^

The emergence of the Omicron variant has led to steep increases in infections and hospitalizations worldwide,^12^ causing significant health losses and disruptions to daily life, commerce and travel, as well as severe stress on the healthcare sector dealing with a high volume of patients in many places across the globe. While Omicron appears to be less severe than other variants,^13^ it is also more transmissible and more able to escape both existing immunity garnered by previous infections and that conferred through vaccination.^14^

Previous literature has discussed the feasibility and costs of global vaccination for COVID-19,^15,16^ placing the total cost somewhere between $25 billion and $50 billion.^17,18^ However, the dangers of not vaccinating come at a potential worldwide cost of trillions of dollars.^17,19^ WHO estimates a delivery cost per vaccine dose of about $1.66, which together with vaccine production costs comes to a cost of about $5 per dose in arm.^15^ Even when incorporating higher vaccine prices as offered by manufacturers, the cost per dose delivered stays below $10.

This analysis aimed to estimate the number of deaths that could be averted via rapid scale up of vaccination in LIC/LMIC around the world, analyzing vaccination scenarios involving either two or three doses of mRNA vaccines. In addition, using estimates of the total cost of vaccine manufacturing and delivery, we evaluated the cost-per-death averted in LIC/LMIC. Our analysis provides lower and upper bounds on estimates of deaths averted under a broad range of assumptions and parameter values. Our study uses mRNA vaccines as the intervention, given that estimates for the costs of their rapid global scale-up are available,^15^ the World Health Organization is already setting up an mRNA production hub,^16^ and they are among the most effective vaccines against COVID-19 to date, particularly against the Omicron variant.^14,20^

## Methods

### Analytic Approach

We developed a model to assess potential deaths averted, total vaccination costs, and cost per death averted assuming vaccination scale up to 100% coverage in LIC/LMIC. We assumed the main COVID-19 variant to be Omicron, which leads to less severe disease than the prior Delta variant but has greater transmissibility and immune escape. The model assumed that all unvaccinated and previously uninfected individuals would be infected with COVID-19 within the year, and some percentage of those infected would die based on infection fatality ratios (IFRs) estimated for the Omicron variant for spike naïve, previously infected, and vaccinated individuals.

Our analysis of avertable deaths proceeded in three steps. First, we used mortality data (both estimates of excess mortality and reported COVID-19 deaths) over the last 22 months to estimate cumulative infections of COVID-19 across LIC/LMIC in all regions. Second, we used the number of previously infected people per country, the number of already vaccinated people per country, and the IFR to estimate a “number of avertable deaths” for future death from COVID-19, assuming those who are previously vaccinated and those who have previous immunity are protected at least in part. Third, we predicted the number of deaths that could be prevented based on estimated vaccine effectiveness.

We evaluated two potential dosage scenarios, one which estimates cost and effect of global vaccination with two doses of mRNA vaccine (“two-dose scenario”), and one with three doses of mRNA vaccine, the primary full course plus an additional booster (“three-dose scenario”). The two-dose scenario assumes that everyone who is unvaccinated at the start of the simulation will receive two doses, and those with one vaccine dose will receive an additional dose. The three-dose scenario calculation assumes those who are unvaccinated will receive three doses, those with two doses will receive one additional dose, and those with one dose will receive two additional doses. For cost calculations, we multiplied the number of doses needed for full population vaccination under these dosage scenarios by the cost per dose.

We estimated the number previously infected with COVID-19 in each region using estimates of regional COVID-19-related mortality and IFR:

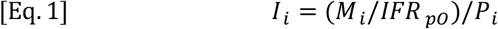

*I*_*i*_ is the proportion of the population in region *i* with previous COVID-19 infection, *M*_*i*_ is the number of deaths attributed to COVID-19 since March 2020, *IFR*_*pO*_ is the infection fatality rate of COVID-19 pre-Omicron, and *P*_*i*_ is the population of region i.

As mortality due to COVID-19 is not well-measured in most LIC/LMIC, we estimate *M*_*i*_in two ways. One method uses reported COVID-19 deaths as the COVID-19 mortality estimate, with the understanding that this would likely underestimate mortality due to COVID-19.

Alternatively, we use excess deaths estimates as the indicator of COVID-19 mortality.^11^ The modeled estimates of excess death have wide confidence intervals, and therefore we use both the median estimate and the 95% confidence limits in our analyses to reflect this uncertainty.

We estimated the number of potentially avertible deaths (*D*_*i,d*_) from COVID-19 based on prior immunity and prior vaccination in each region with the two-dose and three-dose scenario:

For two-dose scenario:

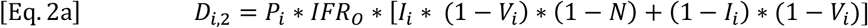

For three-dose scenario:

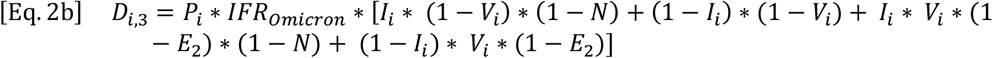

*I*_*i*_ is calculated in equation 1, *IFR*_*O*_ is the infection fatality rate of the Omicron variant of COVID-19, *V*_*i*_ is the proportion of the population in region *i* previously vaccinated with two doses of mRNA vaccine, *N* is the protection against mortality conferred by natural immunity, and *E*_*2*_ is the vaccine effectiveness against mortality for two doses of mRNA vaccine.

We then estimated the number of deaths averted (*A*_*I,d*_):

For two-dose scenario:

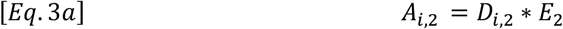

For three-dose scenario:

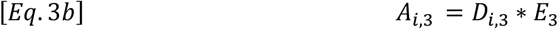

Where *E*_*3*_ is the vaccine effectiveness against mortality from three doses of vaccine.

We estimate the total number of vaccine doses necessary to fully vaccinate a region (*X*_*i,d*_):

For two-dose scenario:

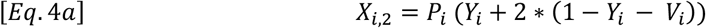

For three-dose scenario:

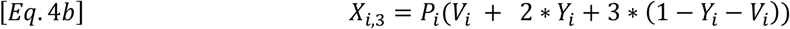

*Y*_*i*_ is the proportion of the population of region *i* vaccinated with only one dose of vaccine.

We estimated total cost of vaccination for each region (*T*_*i,d*_):

For two-dose scenario:

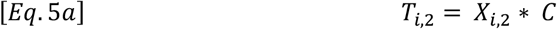

For three-dose scenario:

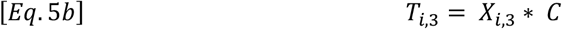

*X*_*i*_ comes from equations 4a and 4b, and *C* was the cost per vaccine dose. Finally, we estimated cost per death averted (*Z*_*i,d*_):

For two-dose scenario:

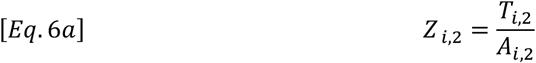

For three-dose scenario:

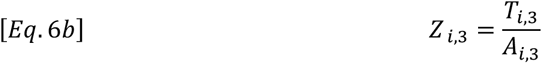

*T*_*i,d*_ comes from equations 5a and 5b, and *A*_*i*.*d*_ comes from equations 3a and 3b.

### Parameter Values

Data on population by region and number of vaccinations came from Our World in Data and WHO (Table 1).^2,12^ Data on reported COVID-19 mortality and excess mortality by region for 2020 and 2021 came from the Institute for Health Metrics and Evaluation and from *The Economist*.^11,21^

**Table 1.** Population characteristics by region, low income and lower-middle income countries.

|  | **Total** | **Sub-Saharan**<br>**Africa** | **Europe and**<br>**Central Asia** | **Latin**<br>**America and**<br>**the**<br>**Caribbean** | **South Asia** | **East Asia and**<br>**the South**<br>**Pacific** | **Middle East and**<br>**North Africa** |
| --- | --- | --- | --- | --- | --- | --- | --- |
| **Characteristics** |  |  |  |  |  |  |  |
| Population<br>(millions) | 3,669 | 1,092 | 98 | 46 | 1,876 | 303 | 253 |
| % Vaccinated | 37.4% | 7.3% | 32.6% | 40.9% | 54.3% | 50.7% | 27.3% |
| % Fully<br>Vaccinated | 24.3% | 3.7% | 22.9% | 32.0% | 34.2% | 39.5% | 20.9% |
| Number<br>Reported<br>COVID-19<br>Deaths<br>(thousands) | 945 | 54 | 110 | 35 | 566 | 103 | 77 |
| Number<br>Estimated<br>Excess Deaths<br>from COVID-19 (95% CI)<br>(thousands) | 9,309<br>(2,612-13,782) | 1,393 (0-2,240) | 272 (227-313) | 154 (89-185) | 6,495 (1,787-9,554) | 438 (231-700) | 556 (317-791) |

Countries were assigned to regions based on World Bank designations.^1^ Only low-income (LIC) and lower-middle income countries (LMIC) were included in the main analysis. Population characteristics for each are shown in Table 1.

We extracted estimates of the infection fatality rate (IFR, for pre-Omicron and Omicron), natural immunity protection against mortality following past COVID-19 infection, vaccine effectiveness against mortality, and cost per dose of vaccine administered from the published literature (Table 2).

**Table 2.** Model parameter estimates and ranges.

|  | Base case<br>parameters | Sensitivity Analysis | Source |
| --- | --- | --- | --- |
| Parameter |  |  |  |
| Infection fatality ratio (IFR) for pre-Omicron<br>COVID-19 | 5/1,000 |  | Levin et al.<sup>22</sup> |
| Infection fatality ratio (IFR) for Omicron variant of<br>COVID-19 | 1/1,000 | 5/10,000<br>5/1,000 | Levin et al.<sup>22</sup><br>Lewnard et<br>al.<sup>24</sup><br>Abdullah et<br>al.<sup>13</sup> |
| Natural immunity protection against COVID-19<br>mortality | 80% |  | Bozio et al.<sup>23</sup> |
| Vaccine effectiveness against COVID-19 mortality,<br>two dose | 80% | 80%<br>95% | Sheikh et al.<sup>30</sup> |
| Vaccine effectiveness against COVID-19 mortality,<br>two dose + booster | 90% | 90%<br>99% | Sheikh et al.<sup>30</sup> |
| Cost per vaccine dose administered | \$7 | \$5-\$10 | Public<br>Citizen<sup>15</sup><br>WHO<sup>16</sup> |

For the base case analysis, we chose realistic parameter values with regards to the cost and benefits of scaling global COVID-19 vaccination. We set the COVID-19 Omicron variant IFR as one fifth of the pre-Omicron estimate (1 in 1,000 vs 5 in 1,000),^13,22^ natural immunity protection against mortality as 80%,^23^ and vaccine effectiveness against mortality as 80% for the two-dose scenario and 90% for the three-dose scenario (Table 1). Production and delivery of an mRNA vaccine dose was estimated to cost US$7, the halfway point between relevant estimates of $5 to $10.^15,16^

### Sensitivity Analysis

Given the uncertainty surrounding several model parameters, we sought to assess the robustness of our conclusions under a wider range of assumptions about the values of our parameters alone and in combination. We therefore conducted a sensitivity analysis assessing the number of deaths averted and the cost-per-death averted while varying IFR, vaccine effectiveness, and vaccine cost. We ranged IFR from a “high” estimate of 5/1,000 consistent with pre-Omicron levels,^22^ to a “low” estimate of 5/10,000, consistent with mortality reduction observed for Omicron in the US.^24^ We ranged vaccine effectiveness against mortality for the two-dose scenario from 80% to 95%, and for the three-dose scenario from 90% to 99%, given the uncertainty of vaccine effectiveness against Omicron as well as potential future variants.^25^ Vaccine cost was ranged from $5 to $10 per dose (Table 1).

As a scenario analysis, we examined the value of vaccines under imperfect uptake. For this analysis, avertible deaths were reduced from 100% to 75%, consistent with vaccine uptake observed in the United States. Cost for vaccine production was kept at 100%, with the assumption that enough vaccine is produced for full vaccination but uptake is reduced.

We also assessed the potential benefit of vaccination in a scenario of high prior infection with COVID-19, since Omicron may spread through the world faster than vaccination can take place. This sensitivity analysis assumed that 100% of the population in LIC/LMIC would be previously infected by COVID-19 prior to vaccination, such that mortality risk is entirely among those with partial natural protection.

## Results

### Base Case Analysis

For the two-dose scenario with base case parameter estimates, using excess mortality to estimate prior infections and number at risk, scaling up vaccination to provide two doses of mRNA vaccine to everyone in LIC/LMIC would cost $35.5 billion and avert 1.3 million deaths from COVID-19, at a cost of $26,900 per death averted (Table 3). Scaling up vaccination to provide three doses of mRNA vaccine to everyone in LIC/LMIC would cost $61.2 billion and avert 1.5 million deaths from COVID-19 at a cost of $40,800 per death averted (Table 3).

**Table 3.** Model results for total cost of global vaccination of low and lower-middle income countries (in billions US$), total deaths averted (in millions), and cost per death averted (US$). The model assumes an IFR for pre-Omicron COVID: 5/1000, IFR for Omicron: 1/1,000, cost per dose $7, natural immunity protection against mortality 80%, vaccine effectiveness against mortality with two doses: 80%, vaccine effectiveness against mortality with three doses: 90%.

|  | Two Dose Scenario | Three Dose Scenario |
| --- | --- | --- |
| **Total cost (US\$, billion)** | \$35.52 | \$61.21 |
| **Total vaccine doses needed (billion)** | 5.07 | 8.74 |
| **Total deaths averted (million)** |  |  |
| Using reported COVID-19 deaths to calculate prior infection | 2.14 | 2.40 |
| Using excess mortality estimates to calculate prior infection (and 95% CI)<sub>1</sub> | 1.32 (0.87-1.97) | 1.50 (1.02-2.22) |
| **Cost per death averted (US\$)** | | |
| Using reported COVID-19 deaths to calculate prior infection | \$16,700 | \$25,500 |
| Using excess mortality estimates to calculate prior infection (and 95% CI)<sub>1</sub> | \$26,900 (\$18,000-\$40,000) | \$40,800 (\$27,600-\$59,900) |

1. 95% CI from excess mortality calculations from The Economist<sup>11</sup>

### Sensitivity Analysis

We considered a wide range of values for IFR and vaccine effectiveness (Table 2). For the two-dose scenario, the cost-per-death averted ranged from $4,500 when IFR was 5/1,000 and vaccine effectiveness against mortality was 95%, to $53,800 when IFR was 5/10,000 and vaccine effectiveness against mortality was 80% (Figure 1). Deaths averted with the same parameter estimates ranged from 7.8 million to 0.7 million, respectively (Figure 2).

**Figure 1.**
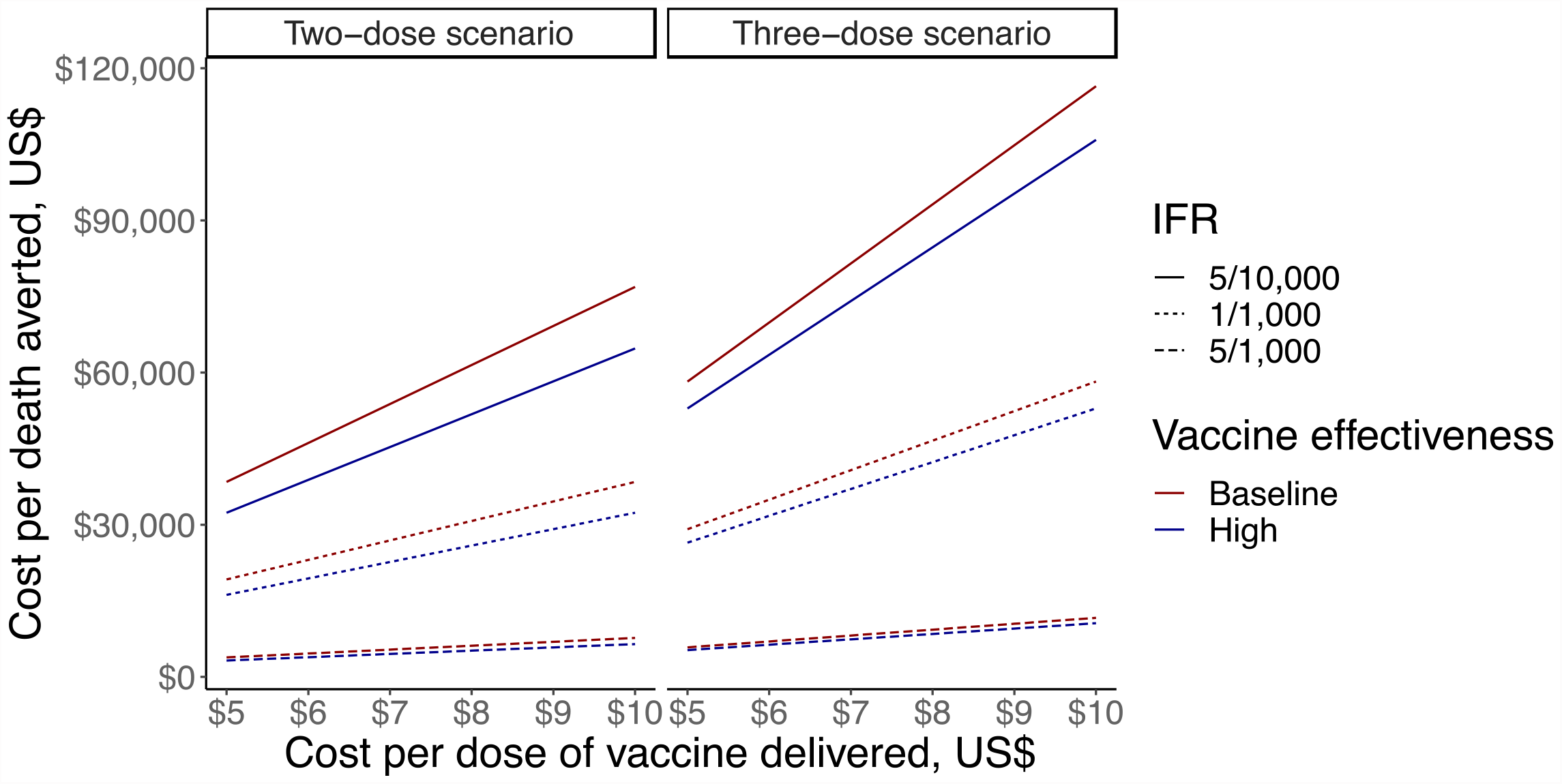
Sensitivity analysis looking at cost-per-death averted of vaccination in LIC/LMIC, ranging cost per vaccine dose, IFR and vaccine effectiveness against mortality in the two-dose scenario (first panel) and three-dose scenario (second panel). The y-axis shows cost-per-death averted in US$, the x-axis shows cost per dose of vaccine in US$. Solid lines show IFR of 5/10,000, dotted line shows IFR of 1/10,000, and dashed lines show IFR of 5/1,000. Dark red lines show baseline vaccine effectiveness (80% in two-dose scenario and 90% in three-dose scenario), and dark blue lines show high vaccine effectiveness (95% in two-dose scenario and 99% in three-dose scenario).

**Figure 2.**
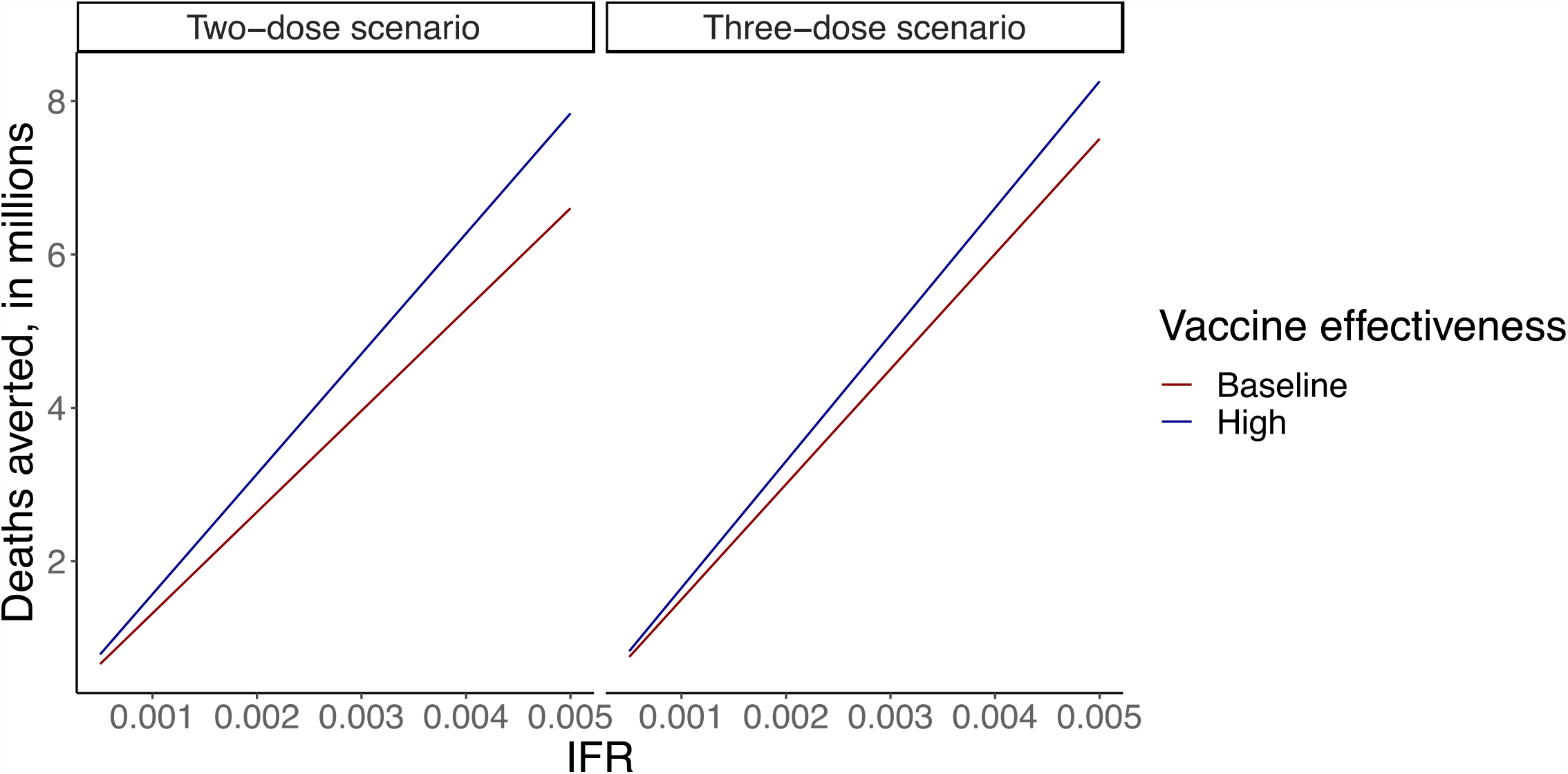
Sensitivity analysis looking at deaths averted of vaccination LIC/LMIC, ranging IFR and vaccine effectiveness against mortality in the two-dose scenario (first panel) and three-dose scenario (second panel). The y-axis shows deaths averted, in millions. The x-axis shows IFR. Dark red lines show baseline vaccine effectiveness (80% in two-dose scenario and 90% in three-dose scenario), and dark blue lines show high vaccine effectiveness (95% in two-dose scenario and 99% in three-dose scenario).

Considering an IFR of 1/1,000 and a vaccine effectiveness of 80% with the two-dose scenario, varying the cost-per-dose from $5 to $10 led to a range of cost-per-death estimates of $19,200 with a $5 cost-per-dose to $38,400 with a $10 cost-per-dose (Figure 1).

Examining the implication of this parameter uncertainty for the three-dose scenario, the cost-per-death averted ranged from $7,400 when IFR was 5/1,000 and vaccine effectiveness against mortality was 99% to $81,500 when IFR was 5/10,000 and vaccine effectiveness against mortality was 90% (Figure 1). Deaths averted with the same parameter estimates ranged from 8.3 million to 0.8 million, respectively (Figure 2).

Considering an IFR of 1/1,000 and a vaccine effectiveness of 80% with the three-dose scenario, varying the cost-per-dose from $5 to $10 led to a range of cost-per-death estimates of $29,100 with a $5 cost-per-dose to $58,200 with a $10 cost-per-dose (Figure 1).

### Vaccine Hesitancy Analysis

For the two-dose scenario with base case parameter estimates, using excess mortality to estimate prior infections and number at risk, reducing vaccine uptake to 75% but keeping cost at 100% would avert 1.0 million deaths at a cost of $25,900 per death averted. When considering a wide range of values for IFR and vaccine effectiveness, cost-per-death averted ranged from $7,200 when IFR was 5/1,000 to $71,700 when IFR was 5/10,000. Deaths averted with the same parameter estimates ranged from 5.0 million to 0.5 million, respectively.

For the three-dose scenario with base case parameter estimates, using excess mortality to estimate prior infections and number at risk, reducing vaccine uptake to 75% but keeping cost at 100% would avert 1.2 million deaths at a cost of $53,400 per death averted. When considering a wide range of values for IFR and vaccine effectiveness, cost-per-death averted ranged from $10,700 when IFR was 5/1,000 to $107,700 when IFR was 5/10,000. Deaths averted with the same parameter estimates ranged from 4.7 million to 0.6 million, respectively.

### All Previously Infected Scenario

We finally considered a scenario in which Omicron infects 100% of LIC/LMIC population before vaccination is available, and all mortality risk occurs through re-infection. In this instance, the three-dose scenario with natural immunity protection against mortality of 80% and base case parameters leads to a cost-per-death averted at $115,000, with 0.5 million deaths averted.

When varying IFR and vaccine effectiveness against mortality, with a cost-per-dose of $7 cost-per-death averted ranged from $20,900 with an IFR of 5/1,000 and vaccine effectiveness against mortality of 99% and $230,000 with and IFR of 5/10,000 and vaccine effectiveness against mortality of 90%. Deaths averted with the same parameter estimates ranged from 2.9 million to 0.3 million, respectively.

## Discussion

Our analysis suggests that scale-up of vaccination in LIC/LMIC is achievable at justifiable cost, with cost-per-death averted estimates between $7,400 and $81,500 for three doses of mRNA vaccine with a $7 cost per dose. While the estimated cost-per-death averted in LIC/LMIC varies with estimated infection fatality ratio and vaccine effectiveness against a given COVID-19 variant, all estimates of cost-per-death averted in our analysis are well below the commonly used estimates for the value of a statistical life (VSL), with accepted estimates ranging from €3.7 million in Sweden to AUS$7.3 million in Australia.^26,27^ Even though cost to avert a death rises at lower COVID-19 IFR, lower vaccine effectiveness, and higher cost per dose, it is a fraction of the US 2022 VSL estimates of $4.9 million - $15.9 million.^28^

Our current analysis focuses on the Omicron variant of COVID-19, a more transmissible but clinically less severe variant than those previously seen. When applying an infection fatality rate similar to that seen in previous variants of COVID-19, the number of deaths averted would rise to over 10 million and the cost-per-death averted would drop below $10,000. If future variants of COVID-19 are more severe than Omicron, the imperative to vaccinate becomes more acute.

Our model does not account for demographic differences among regions, including age structure and the prevalence of comorbid conditions which could affect COVID-19 mortality. Our analysis also did not include limitations on the capacity of health systems, which could affect mortality beyond what we capture with IFR. In addition, we do not attempt to model disease transmission dynamics, or include a temporal component in the model, so we assume anyone without immunity from vaccine or natural protection as of December 13, 2021 retains their risk of COVID-19 mortality through 2022. This is a major limitation given the speed with which the Omicron variant of COVID-19 has been spreading; however, our analysis considering vaccination scale-up after the whole population has been previously infected still estimates a cost-per-death averted of $115,000. Finally, our main analysis uses reported deaths or excess mortality estimates to estimate rates of prior infection with COVID-19, which are inexact measures especially for the regions of most concern. Nevertheless, they are the best available estimates to date.

Experts have proposed several plans to rapidly expand global COVID-19 vaccine production and delivery.^15,17,18^ However, the international community has not yet invested the resources required nor displayed the political commitment necessary to scale global vaccination. As a result, 86 countries did not reach the World Health Organization’s target of vaccinating 40 percent of their populations by the end of 2021.^29^ This analysis shows that global vaccination can be undertaken for a fraction of the trillions of US$ already spent on global COVID-19 response, and would avert deaths for a cost well below recognized VSL estimates. Whether the investment of US and other donor nations’ resources for global COVID-19 vaccination are worth spending for this endeavor depends on how we value the lives in low and lower middle-income countries.

## Data Availability

This study used only publicly available data sources.

## Notes

### Competing Interest Statement

The authors have declared no competing interest.

### Funding Statement

Funding: MCF gratefully acknowledges funding from the National Institutes of Health (5K01AI141576). AB and JAS were supported by the Centers for Disease Control and Prevention though the Council of State and Territorial Epidemiologists (NU38OT000297-02; AB, JAS).

